# L-Dopa-induced changes in aperiodic bursts dynamics relate to individual clinical improvement in Parkinson’s disease

**DOI:** 10.1101/2024.06.14.24308683

**Authors:** Hasnae Agouram, Matteo Neri, Marianna Angiolelli, Damien Depannemaecker, Jyotika Bahuguna, Antoine Schwey, Jean Régis, Romain Carron, Alexandre Eusebio, Nicole Malfait, Emmanuel Daucé, Pierpaolo Sorrentino

## Abstract

Parkinson’s disease (PD) is a neurodegenerative disease primarily characterized by severe motor symptoms that can be transiently relieved by medication (e.g. levodopa). These symptoms are mirrored by widespread alterations of neuronal activities across the whole brain, whose characteristics at the large scale level are still poorly understood. To address this issue, we measured resting state activities of 11 PD patients using DBS contacts in the subthalamic nucleus (STN) and EEG electrodes over motor areas. Data were recorded for each patient before (OFF-condition) and after (ON-condition) levodopa administration. Neuronal avalanches, i.e. brief bursts of activity with widespread propagation, were detected and quantified on both types of contacts, and used to characterize differences in both conditions. Of particular interest, we noted a larger number of shorter and smaller avalanches in the OFF-condition, and a lesser number of wider and longer avalanches in the ON-condition. This difference turned out to be statistically significant at the group level. Then, we computed the avalanche transition matrices (ATM) to track the contact-wise patterns of avalanche spread. We observed a higher probability that an avalanche would spread within and between STN and motor cortex in the ON-state, with highly significant differences at the group level. Furthermore, we discovered that the increase in overall propagation of avalanches was correlated to clinical improvement after levodopa administration. Our study offers the initial cross-modality assessment of aperiodic activities in PD patients, including levodopa’s effects on cross-regional aperiodic bursts at the individual level, suggesting potential biomarkers for PD electrophysiological alterations.

**Significance Statement:** Our research focuses on levodopa’s effects on large-scale dynamics in PD using a novel approach involving aperiodic bursts (i.e., neuronal avalanches). To achieve this, we measured resting-state activities of 11 PD patients using DBS contacts in the STN and EEG electrodes placed bilaterally over the motor areas, both before and after levodopa administration. Unlike most studies on beta frequency (13–30 Hz) activities, we examined dynamics through aperiodic bursts across temporal and spatial scales. Instead of focusing on global properties, we tracked the spatial spread of neuronal avalanches across the brain. Our study provides the first assessment of levodopa’s effects on cross-regional aperiodic bursts within and between the STN and motor cortex, and suggests potential electrophysiological biomarkers for PD.

## Introduction

Parkinson’s disease (PD) is a chronic and progressive neurological disorder that affects movement, with key motor symptoms including tremor, rigidity, bradykinesia, and postural instability (Jankovic, 2008; Poewe et al., 2017). Additionally, the clinical picture also encompasses several non-motor symptoms such as depression, sleep disturbances, and dementia (Hou and Lai, 2007; Armstrong and Okun, 2020). The pathological hallmark of PD involves the destruction of dopaminergic neurons in the pars compacta of the substantia nigra (Bernheimer et al., 1973; Jucker and Walker, 2013; Armstrong and Okun, 2020). Previous work showed that substantia nigra malfunction affects the entire brain, which is mirrored by widespread alterations in neuronal activities in PD patients (Braak et al., 2004; Weingarten et al., 2015; Oswal et al., 2016a; Kim et al., 2017; Ma et al., 2017). In particular, interactions within the basal ganglia-thalamocortical circuit have been central in characterizing and studying the mechanisms underpinning this disease (Marreiros et al., 2013). To investigate these, we leveraged a unique dataset with simultaneous bilateral recordings: EEG in the motor cortex and deep stimulation electrodes in the subthalamic nuclei (STN). Such an approach may prove beneficial in bridging the gap between local pathological changes (e.g. within the cortical-BG-thalamic loop) and the corresponding clinical manifestations in PD (by elucidating how local alterations reverberate across the whole brain).

Classically, large-scale interactions in PD have been understood in terms of periodic oscillations and their modulation (i.e. beta bursts) (Brown et al., 2004; Kuhn et al., 2008; Oswal et al., 2016b; Trager et al., 2016; Tinkhauser et al., 2017; Wang et al., 2018). More recently, a new perspective has focused on the scale-free properties of large-scale activities (Beggs and Plenz, 2003; Shriki et al., 2013). Aperiodic, spontaneous bursts spreading across a range of spatial and temporal scales (“neuronal avalanches”) have been consistently observed across different imaging modalities and spatio-temporal scales (Tagliazucchi et al., 2012; Palva et al., 2013; Priesemann et al., 2013). Neuronal avalanches, manifesting as bursts of activations spreading across multiple brain signals, characterize the large-scale interactions across multiple brain regions, and constitute a marker of physiological states, for example allowing the characterization of conditions such as sleep and resting wakefulness (Priesemann et al., 2013; Scarpetta et al., 2023), or speech and music listening (Neri et al., 2023). Furthermore, avalanche dynamics were shown to be altered in PD patients, and proportionally to clinical impairment (Sorrentino et al., 2021a).

While avalanches have been classically analyzed in terms of their global statistical properties (e.g. the distribution of their sizes, durations, flexibility, etc.), a more recent approach focused on the topography and topology of such bursts across the brain. To this end, the avalanche transition matrices (ATMs) have been developed to track from where to where avalanches spread on average across the brain. As a result, it became evident that the spatial and temporal propagation patterns differ between healthy state and pathology (Seshadri et al., 2018; Rucco et al., 2020; Sorrentino et al., 2021a; Polverino et al., 2022; Duma et al., 2023).

In the present work, we deploy for the first time the analysis of avalanches in a cross-modality setting, under the hypothesis that the administration of levodopa changes the spreading of aperiodic bursts of activities between the motor cortex and the subthalamic nuclei . To achieve this, we recorded the resting state activities of 11 PD patients in two conditions: ON-levodopa and OFF-levodopa medication. We hypothesize that the changes induced by levodopa on large-scale activities can be understood in terms of changes in the topology of scale-free perturbations, manifesting as different propagation patterns and, more generally, different statistical properties of large-scale perturbations. As such, avalanches would present different statistical features and propagate differently according to the patient’s medication state which, in turn, would be related to clinical symptoms. Hence, the ATMs might be seen as a potential marker to track both the medication effects and motor improvement at the individual level.

## Materials and Methods

### Participants, Surgery, Experiments, and Data Collection

The study involved 11 patients (22 hemispheres) diagnosed with advanced Parkinson’s disease who underwent deep brain stimulation (DBS) surgery targeting the subthalamic nucleus (STN). Recordings were taken using Deep Brain Stimulation leads and EEG electrodes placed bilaterally over the motor areas. The recordings were performed under two conditions: OFF-levodopa medication (before its administration) and ON-levodopa medication (after its administration).

The clinical information of the patients is reported in the Extended Data section (Table1). The local ethics committee approved the study, and all participants provided written informed consent. Bilateral DBS surgery was performed using Medtronic 3389 DBS leads. Intra-operative micro-recordings and macro-stimulation during surgery were utilized for accurate lead placement, and postoperative clinical assessments, supported by the fusion of preoperative MRI and postoperative CT scans for validation, were performed. The experimental procedure involved temporary externalization of DBS electrodes before connecting them to the implantable pulse generator, usually five days later. Patients abstained from taking dopaminergic medication the night before recordings. Brain signals were recorded for 2–3 minutes with patients at rest, seated comfortably. Simultaneous recordings of local field potentials (LFPs) and scalp EEG signals were obtained, including 8 LFP contacts for the subthalamic nuclei (4 for the Left STN and 4 for the Right STN), and 6 EEG channels in the bilateral motor areas (F3, Fz, F4, C3, C4, Cz). Spike artifacts were minimized and removed using the Spike2 Software. Data was imported into Matlab, with signal durations ranging from 119.2 s to 155.5 s and a mean duration of 127.2 s ± 2.5. The manuscript utilizes previously acquired data. See (Tinkhauser et al., 2018) for more information.

### Data preprocessing

Signal preprocessing was performed using the Fieldtrip toolbox (Oostenveld et al., 2011). The continuous EEG signal was first high-pass filtered at 1.3 Hz with a Hamming window, using a ‘two-pass’ direction, and a Butterworth filter. Subsequently, the data was down-sampled to 512 Hz and epoched into 4-second segments. The signal underwent visual inspection to remove noisy segments.

### Estimation of neuronal avalanches

Neuronal avalanches were estimated by binarizing z-scored activities (each brain signal was independently z-scored across time) to identify salient events in neural dynamics that exceeded a threshold of 2 standard deviations (|z| = 2). When a brain signal has a z-score above this threshold, it is classified as active (assigned a value of 1), while all other time points are considered inactive (assigned a value of 0). A neuronal avalanche initiates when, in a sequence of consecutive time bins, at least one channel becomes active (|z| > 2) and concludes when all channels return to an inactive state (Beggs and Plenz, 2003; Shriki et al., 2013).

Neuronal avalanches are characterized by their size, *s*, defined as the number of channels recruited during the avalanche. They can also be characterized by the duration, and the inter-avalanche interval (IAI), defined as the time interval between two consecutive avalanches.

Our prominent interest was to assess the effect of levodopa medication on neuronal avalanches features, such as their number, size, duration, as well as the inter-avalanche interval. To achieve this, we compared the number of avalanches per segment (i.e., the total number of avalanches divided by the number of time segments), as the exact same amount of time segments was not available for the ON-levodopa and the OFF-levodopa conditions in each patient. At the subject level, we used the Mann-Whitney test to assess the difference in the distributions of the number of avalanches across segments between ON and OFF conditions. At the group level, we employed the paired t-test to assess the significance of the number of avalanches per segment in ON-levodopa versus OFF-levodopa conditions.

We also compared the size, duration, and inter-avalanche interval between ON-levodopa and OFF-levodopa conditions at both the subject and group levels. We used the two-sample Kolmogorov-Smirnov (K-S) test to assess the significance of the differences in the distributions of size, duration, and inter-avalanche interval across segments between ON-levodopa and OFF-levodopa conditions. At the group level, we gathered avalanche sizes, durations, and inter-avalanche intervals for all patients in both ON and OFF conditions. Then, we conducted the K-S test to compare the distributions of these features between ON and OFF conditions. The reason behind choosing the K-S test earlier in the manuscript is its lack of assumptions of gaussianity, given the non-linear, fat-tailed nature of avalanche dynamics (Beggs and Plenz, 2003).

### Avalanche Transition Matrix

We calculated an avalanche-specific transition matrix (TM) where rows and columns represent channels. The element (i, j) of this matrix represents the probability that channel j was active at time t + 1, given that channel i was active at time t (the ij-th entry). Thus, the edge between channels i and j indicates the probability of these two channels being sequentially recruited by an avalanche. For each patient, we obtained an average transition matrix by averaging edge-wise over all avalanches and then symmetrized it (Sorrentino et al., 2021b). Therefore, we obtained two symmetric ATMs per patient, one corresponding to the ON condition and the other to the OFF condition.

### Statistical Analysis

For each patient, we compared the ATMs in two conditions: ON-levodopa medication and OFF-levodopa medication. This comparison provides insight into the probabilities of perturbations spreading across two brain regions. To confirm this statistically, we randomly shuffled the labels of the avalanche-specific transition matrices for each individual. We repeated this procedure 10,000 times, obtaining, for each edge, the distribution of differences under the null hypothesis that the transition matrices would not capture any difference between the two medication conditions. Note that this method does not require normality of the initial distributions. These distributions were then used to identify edges that showed significant differences between the two medication states at the subject level. The p-values were calculated as the proportion of random differences greater than the observed difference. The significance levels were corrected for multiple comparisons across edges using the Benjamini-Hochberg (BH) correction. Following this procedure, we obtained a matrix for each patient containing the edges that exhibited significant differences between the two conditions. After this step, our focus shifted to edges that were consistently significant across patients for both the ON-levodopa medication and OFF-levodopa medication conditions. This same method was previously employed by (Corsi et al., 2023).

We also explored group-level analysis in addition to within-subject analyses. Specifically, we evaluated the differences in avalanche transition matrices (ATMs) for each edge across all patients between the ON-levodopa and OFF-levodopa conditions and assessed significance using the Wilcoxon signed-rank test. To correct for multiple comparisons, we applied the BH correction.

We also conducted additional analyses at the group level to assess whether our results were driven specifically by edges computed between electrodes belonging to the same/different modality (EEG or deep electrodes), we classified the edges into three groups: *STN-STN,* edges connecting two channels both located in the STN (recorded using deep stimulation electrodes), *cortex-cortex,* edges connecting two channels located both in the motor cortex (recorded using EEG), and *STN-cortex*, that is edges connecting one channel located in the STN with one located in the cortex. Next, we compared the average of each group of edges between the ON-levodopa and the OFF-levodopa conditions at the population level. To assess the significance of the differences, we conducted a Wilcoxon signed-rank test. This procedure was chosen to confirm the significance of the differences in the propagation of avalanches between ON-levodopa and OFF-levodopa states in both short-range connections (within the STN, within the motor cortex) and long-range connections (between the STN and the motor cortex) at the group-level, and to assess whether these differences are modality-independent.

### Clinical correlation analysis

To establish a relationship between the change in the overall propagation of avalanches and clinical improvement after levodopa administration, we assessed the relationship between the clinical improvement and the ratio of the average of ATMs ON and OFF levodopa. The clinical improvement was defined as follows (Tinkhauser et al., 2017):

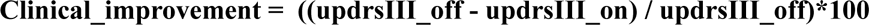

Where ‘updrsIII_off’ and ‘updrsIII_on’ refer to the Unified Parkinson’s Disease Rating Scale Part III before and after medication intake, respectively.

We utilized robust linear regression to obtain more accurate estimates of regression coefficients, reducing the influence of the outliers or deviations from the linear relationship between variables. Specifically, we utilized the “robustfit.m” MATLAB function (Holland and Welsch, 1977), with the bisquare fitting weight function. This function provides coefficient estimates and model statistics as outputs.

We utilized the same technique (i.e. Robust linear regression) to assess the relationship between the ratio of the average ATMs in the ON and the OFF states, computed over three different types of edges (cortex-cortex, STN-cortex, and STN-STN), with the clinical improvement after levodopa administration. In this case, before computing the ratios, we averaged the ATMs ON and the ATMs OFF only considering the edges connecting two channels both located in the cortex for cortex-cortex edges, or edges hinging on the STN and the cortex for STN-cortex edges, and edges connecting two channels both located in the STN for STN-STN edges.

## Results

Neuronal avalanches have been estimated (after signal preprocessing, *see methods*) by binarizing the z-scored activities (each channel was z-scored independently of the others across time), focusing on salient - above threshold (2 standard deviations) - events in neural dynamics. When a brain signal has a z-score above threshold it is set to 1 (*active*); in all the other time points it is set to 0 (*inactive*). A neuronal avalanche starts when at least one channel becomes active, and ends when all channels become inactive. Neuronal avalanches are characterized by their size, *s*, defined as the number of channels recruited during the avalanche; their duration, and the inter-avalanche interval (IAI), defined as the time interval between two consecutive avalanches (Figure 1.A).

**Figure 1.**
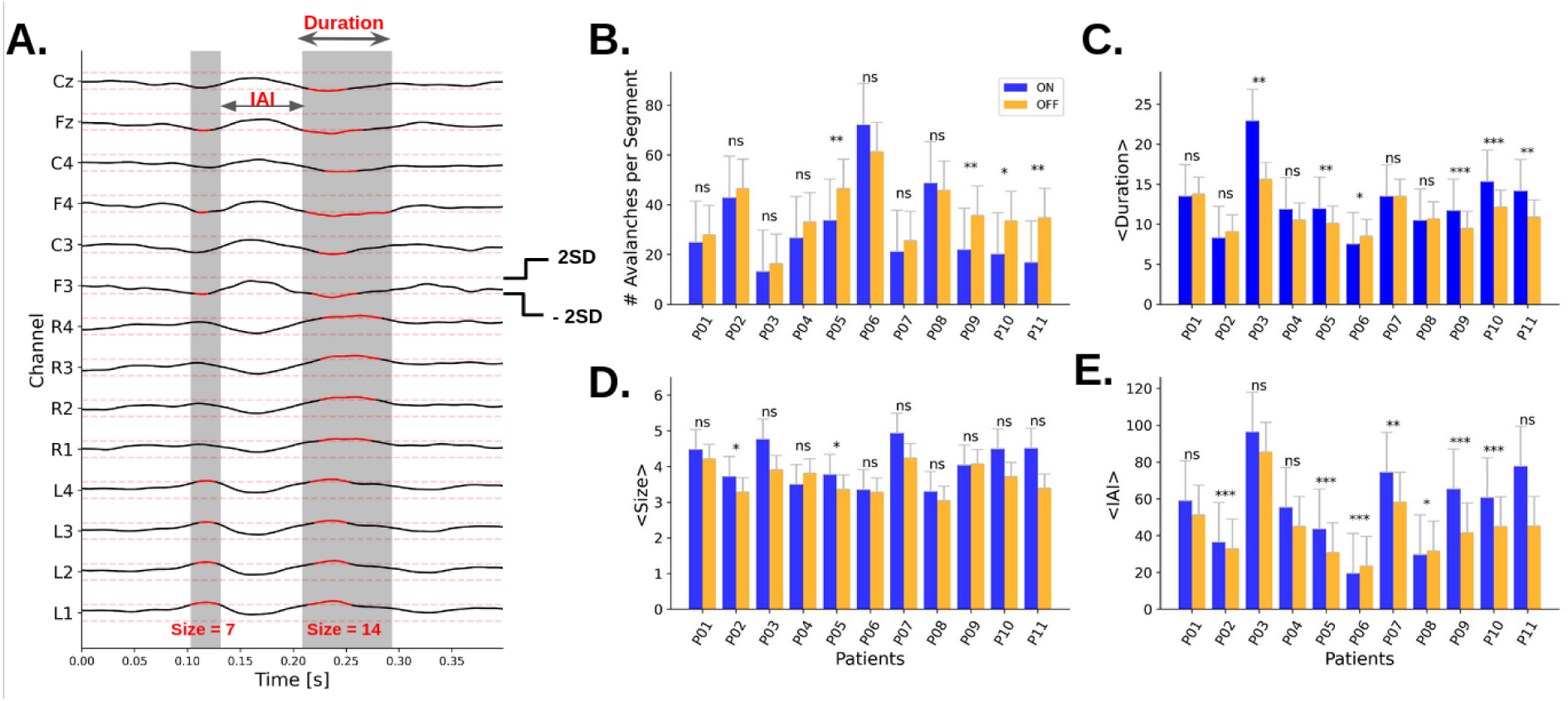
Subject-level analysis of avalanches features. A neuronal avalanche is defined as a continuous sequence of signal excursions beyond the threshold (red thick line) of one or more channels, **(A) Avalanche features illustration** (adapted from (Scarpetta et al., 2023)). The avalanche duration is defined as the total length of an avalanche, while the avalanche size corresponds to the number of recording sites recruited during the avalanche. An avalanche is preceded and followed by intervals during which signals in all channels fall below the threshold (i.e. inter-avalanche intervals). (**B**) **Average number of avalanches per segment, individual comparisons** in the ON-levodopa versus OFF-levodopa states for each patient. The significance of the difference in the distributions of the number of avalanches across segments ON versus OFF was assessed using the Mann-Whitney test. **(C) Avalanche durations (in ms), individual comparisons**. **(D) Avalanche size (# of contacts), individual comparisons. (E) Inter-avalanche intervals (ms), individual comparisons.** For C-D-E, we used the two-sample Kolmogorov-Smirnov (K-S) test to assess the significance of the differences in the distributions of size, duration, and inter-avalanche interval across segments between ON-levodopa and OFF-levodopa conditions. The significance level are indicated as *** : p< 0.001, ** : p<0.01, *: p<0.05, ns = non-significant.

To characterize the two conditions (i.e. ON vs OFF), we first compared the number of avalanches at the subject level (Figure 1.B). We found that avalanches occur more frequently in the OFF-levodopa condition as compared to the ON-levodopa condition in nine patients out of eleven. More specifically, we compared the number of avalanches per segment (i.e., the total number of avalanches divided by the number of time segments), as the exact same amount of time segments was not available for the ON-levodopa and OFF-levodopa conditions in each patient. Segments represent epochs obtained by dividing the continuous resting-state time series into discrete periods. While we reached significance only for four patients out of eleven (Mann-Whitney test), there were more avalanches in the OFF-state as compared to the ON-state in all patients except for two. The group-level analysis (Figure 1-1.A) confirmed a significant difference with a paired t-test (p-value = 0.041). For more details about subject-level analysis, we compared the distributions of the number of avalanches across segments for each patient in the ON-state and the OFF-state (Figure 1-2).

**Figure 2.**
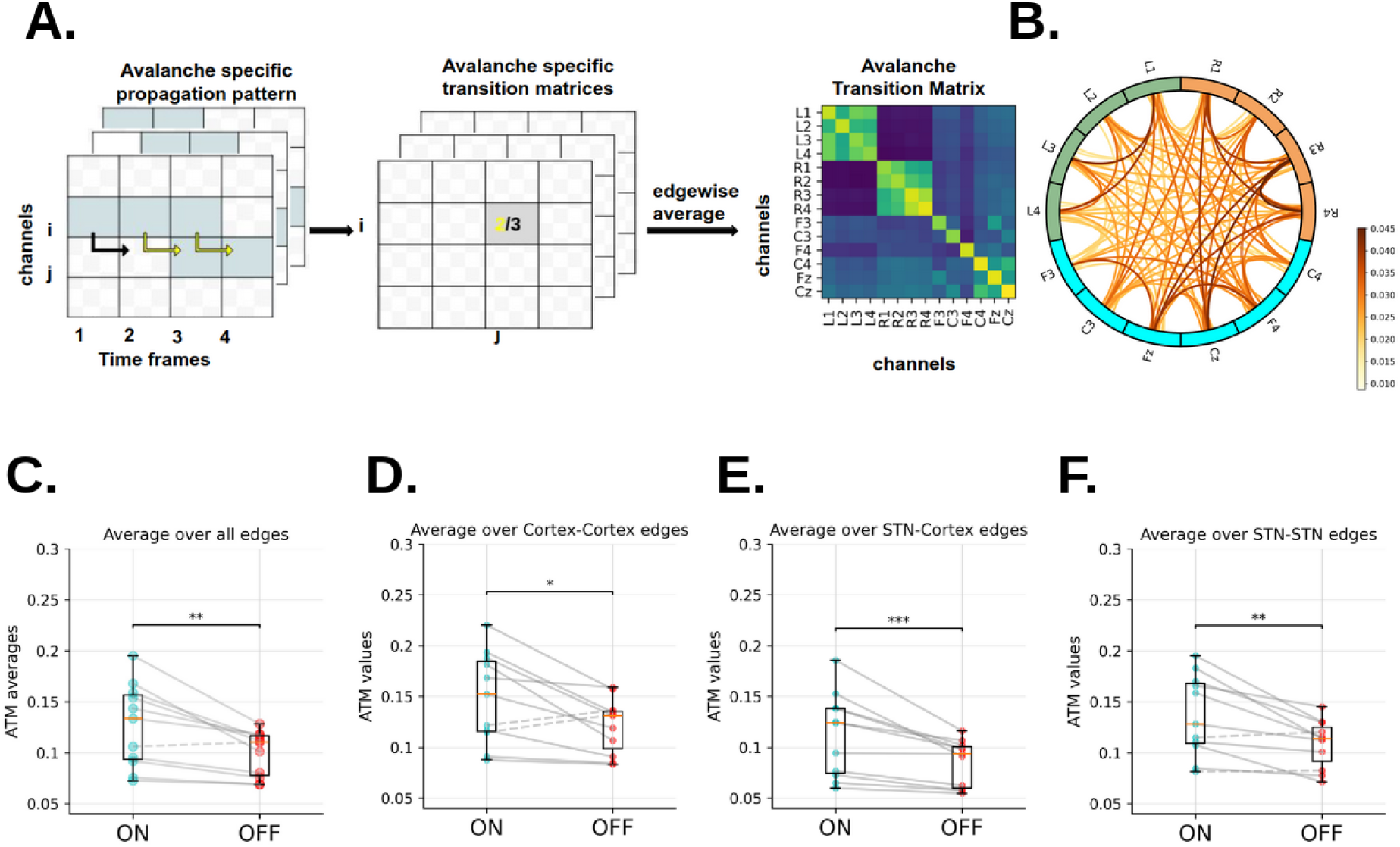
Avalanche Transition Matrix (ATM) in ON-levodopa and OFF-levodopa conditions. **(A) Avalanche Transition Matrix pipeline** The central and left panels are adapted from (Romano et al., 2023). The light blue squares signify that channel i exceeded the threshold three times during the avalanche. Channel j was active following the activation of channel i in just two out of the three cases considered (as shown by the yellow arrows), yielding a probability of 2/3. An individual’s avalanche transition matrix is obtained by averaging over avalanche-specific transition matrices. Right panel: ATMs are structured by channels in rows and columns; the resulting shape being (channels,channels) = (14, 14). **(B) Average edge difference across patients** (circular graph representation) the edge color reports the average difference between the ATMs in ON and OFF-levodopa conditions, across all patients. The left LFP contacts shown in green, right LFP contacts in orange and EEG channels in blue. Regarding the edges, the intensity of the color is proportional to the strength of the difference. In **(C) Group level ATM average comparison** in the ON-levodopa and OFF-levodopa conditions. Each dot represents one patient. The significance of the differences was assessed with the Wilcoxon signed-rank test, yielding a p-value of 0.0029. **(D) Group level Cortico-Cortical edges average comparison, (E) Group level STN-Cortical edges average comparison, (F) Group level STN-STN edges average comparison.** The statistical significance of the differences was tested with the Wilcoxon signed-rank test, resulting in p-values of 0.0322 for Cortex-Cortex edges, 0.0009 for STN-Cortex edges, and 0.0048 for STN-STN edges. The significance level are indicated as *** : p< 0.001, ** : p<0.01, *: p<0.05, ns = non-significant.

Then, we compared the duration (Figure 1.C), size (Figure 1.D), and inter-avalanche interval (IAI) of avalanches (Figure 1.E) at the subject level and we observed a common trend across the majority of the patients: the average size, duration, and inter-avalanche interval (IAI) are larger in the ON-levodopa condition than in the OFF-levodopa condition. While we do not reach statistical significance for all patients utilizing the two-sample Kolmogorov-Smirnov (K-S) test (the reason for choosing the K-S test can be found in the Methods section), the group level (Figure 1-1) confirms a larger size, duration, and inter-avalanche interval in the ON-levodopa condition, with a significant result of the K-S test for the size (p_value=0.048) and inter-avalanche interval (p_value=0.040), while the difference for the durations did not reach statistical significance (p_value=0.069). Overall, our first analysis indicates a higher number of avalanches in the OFF-levodopa state, but larger avalanche size, larger avalanche duration, and longer inter-avalanche intervals in the ON-levodopa state. In conclusion, in the ON-levodopa state, we observe a reduced number of avalanches, yet they exhibit longer durations, greater sizes, and longer intervals between them. For additional details, please refer to Figures 1-3, 1-4, and 1-5 for visualizations of the comparison of the cumulative distribution functions of avalanche sizes, avalanche durations, and inter-avalanche intervals across segments for each patient in the ON-state and OFF-state, along with the results of the Kolmogorov-Smirnov (K-S) test.

**Figure 3.**
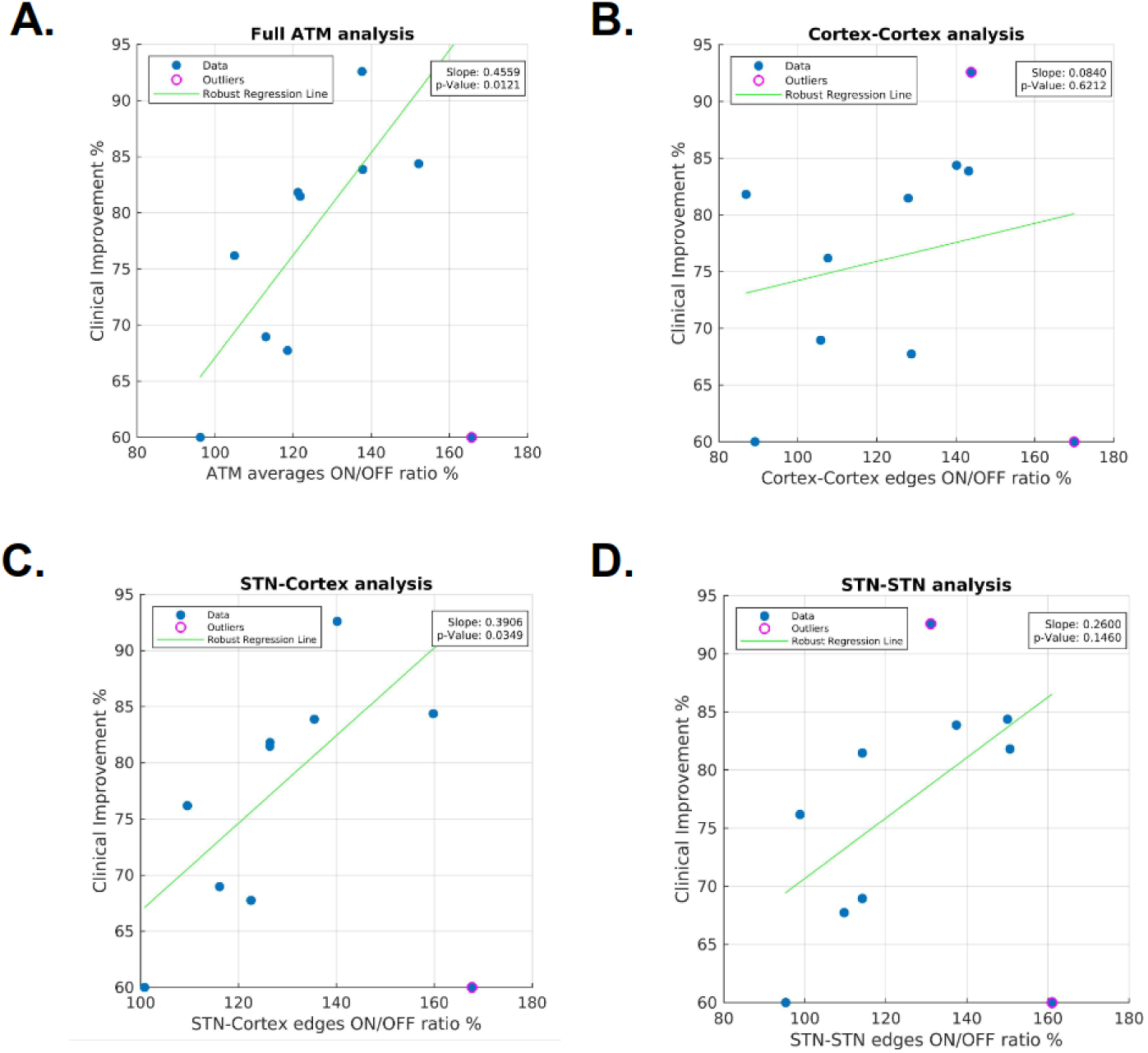
Group level clinical improvement correlation analysis. **(A) ATM averages ON/OFF ratio vs. clinical improvement** Robust linear regression. Each patient is represented as a blue dot, except for one outlier with a pink circle. The regression coefficient was 0.4559, and the p-value was 0.0121. **(B) cortex-cortex edges ON/OFF ratio vs. clinical improvement. (C) STN-cortex edges ON/OFF ratio vs. clinical improvement. (D) STN-STN edges ON/OFF ratio vs. clinical improvement.** Robust linear regression. Each patient is represented as a blue dot, except for the outlier(s) with a pink circle. The regression coefficient and p-values were coef= 0.0840, p= 0.6212 for cortex-cortex edges; coef= 0.3906, p= 0.0349 for STN-cortex edges; and coef= 0.2600, p= 0.1460 for STN-STN edges.

To characterize how avalanches spread throughout the brain, we computed the avalanche transition matrices (Sorrentino et al., 2021c). An avalanche-specific transition matrix was calculated, where element *(i, j)* represents the probability that channel *j* was active at time *t + 1*, given that channel *i* was active at time *t*. For each patient, we obtained an average transition matrix (ATM) (i.e. averaging edge-wise over all avalanches) and then symmetrized it (Figure 2.A). The matrices were symmetrized to help the interpretability of the results and to obtain more reliable estimates. Hence, we obtained two symmetric ATMs for each patient, one for the ON condition and one for the OFF condition, *see methods*.

In other words, ATMs contain values between 0 and 1, which are average probabilities of two nodes to be successively activated by an avalanche during the recording. As observed in Figures 2-1. A and 2-1.B, by averaging all the ATMs in each condition, the probability is higher within each region (i.e. within the left STN, the right STN and the Cortex) than across the areas. However, a clear difference is also visible when we look at the inter-regional links. The darker color in the ON condition indicates that across-region avalanches are more frequent. The average difference between the ATMs (that is, ON-levodopa minus OFF-levodopa) is positive for all the edges across patients (Figure 2.B). This difference is widespread across the regions, and even more pronounced in the right hemisphere. On average, the probability of propagation of avalanches is higher in the ON-levodopa than the OFF-levodopa state, across all pairs of recording sites.

This is confirmed by a statistical analysis. At the group level, the average of ATMs across all edges is significantly higher in the ON-levodopa than in the OFF-levodopa condition (Wilcoxon signed-rank test, p= 0.003) (Figure 2.C). This is true for all the patients of the group except for one. To assess regional effects, we classified the edges into three groups: *STN-STN,* edges connecting two channels both located in the STN (recorded using deep brain stimulation electrodes), *cortex-cortex,* edges connecting two channels located both in the motor cortex (recorded using EEG), and *STN-cortex*, that is edges with one channel located in the STN and the other located in the cortex. We found that the average of each of these was significantly higher during the ON-levodopa than in the OFF-levodopa conditions at the group level (Wilcoxon signed-rank test, STN-STN: p=0.0048; STN-cortex: p=0.0009; cortex-cortex: p=0.0322), see Figure 2.D-E-F.

Those results generally support the global impact of levodopa medication on avalanche dynamics. The statistical difference is more pronounced here, reaching an extremely significant difference (p<0.001) at the level of the STN-cortex edges. This can be interpreted as an enhanced propagation of neuronal avalanches under levodopa medication, this effect being more manifest across regions when considering the subcortical contacts together with the EEG channels. Furthermore, this analysis supports our findings indicating that these are not dependent on modality.

Furthermore, UDPS III scores (*Unified Parkinson’s Disease Rating Scale part III*) were assessed before and after levodopa administration. We thus defined a clinical improvement ratio (*see Methods*), reflecting a percentage of clinical improvement with respect to the baseline score. Then, we assessed the correlation between the clinical improvement and the ratio of the mean ATMs in the ON versus OFF states using robust linear regression *(see Methods)*. A significant positive correlation was found (regression coefficient: 0.4559, p-value: 0.0121), see Figure 3.A, indicating a possible relation between avalanche spread pattern and clinical improvement : as the overall propagation of avalanches increases, the mean clinical improvement after levodopa administration also tends to increase.

We also observed similar results when assessing the relationship of clinical improvement with the ratio between the averages over specific types of edges in the ATM (i.e. cortex-cortex, STN-cortex, and STN-STN) in the ON and OFF states. Our findings indicated a positive correlation for the three types of edges, but it only reached significance for the STN-cortex edges (for cortex-cortex edges, the regression coefficient and p-values were 0.0840 and 0.6212, respectively; for STN-cortex edges, 0.3906 and 0.0349, respectively; and for STN-STN edges, 0.2600 and 0.1460, respectively), see Figure 3.B-C-D. Therefore, clinical improvement appears to relate more specifically to the large-scale coordination of activities, in particular between the STN and the motor cortex.

At a more fine-grained level, we tried to assess whether specific contact-wise avalanche spread differences could be identified within and across patients. In particular, we noticed that while globally the ATMs of the ON-levodopa condition show higher values with respect to the OFF-levodopa condition, there were specific edges and patients in whom we observed the opposite trend. For this reason, we used a permutation test to assess, at the patient level, which edges were significantly stronger in the ON-levodopa with respect to the OFF-levodopa conditions. For each patient, we compared the observed edge-wise difference between the ATM of the ON-levodopa and the OFF-levodopa conditions, with an edge-specific, patient-specific null distribution obtained by iteratively randomizing the labels (ON-levodopa and OFF-levodopa medication) of avalanche-specific transition matrices before averaging (Figure 4.A), *see methods*. In this way, we obtained for each patient, a set of significantly stronger edges (p<0.05, Benjamin-Hochberg corrected across edges) in the ON-levodopa with respect to the OFF-levodopa state, and a set of significantly stronger edges in the OFF-levodopa with respect to the ON-levodopa.

**Figure 4.**
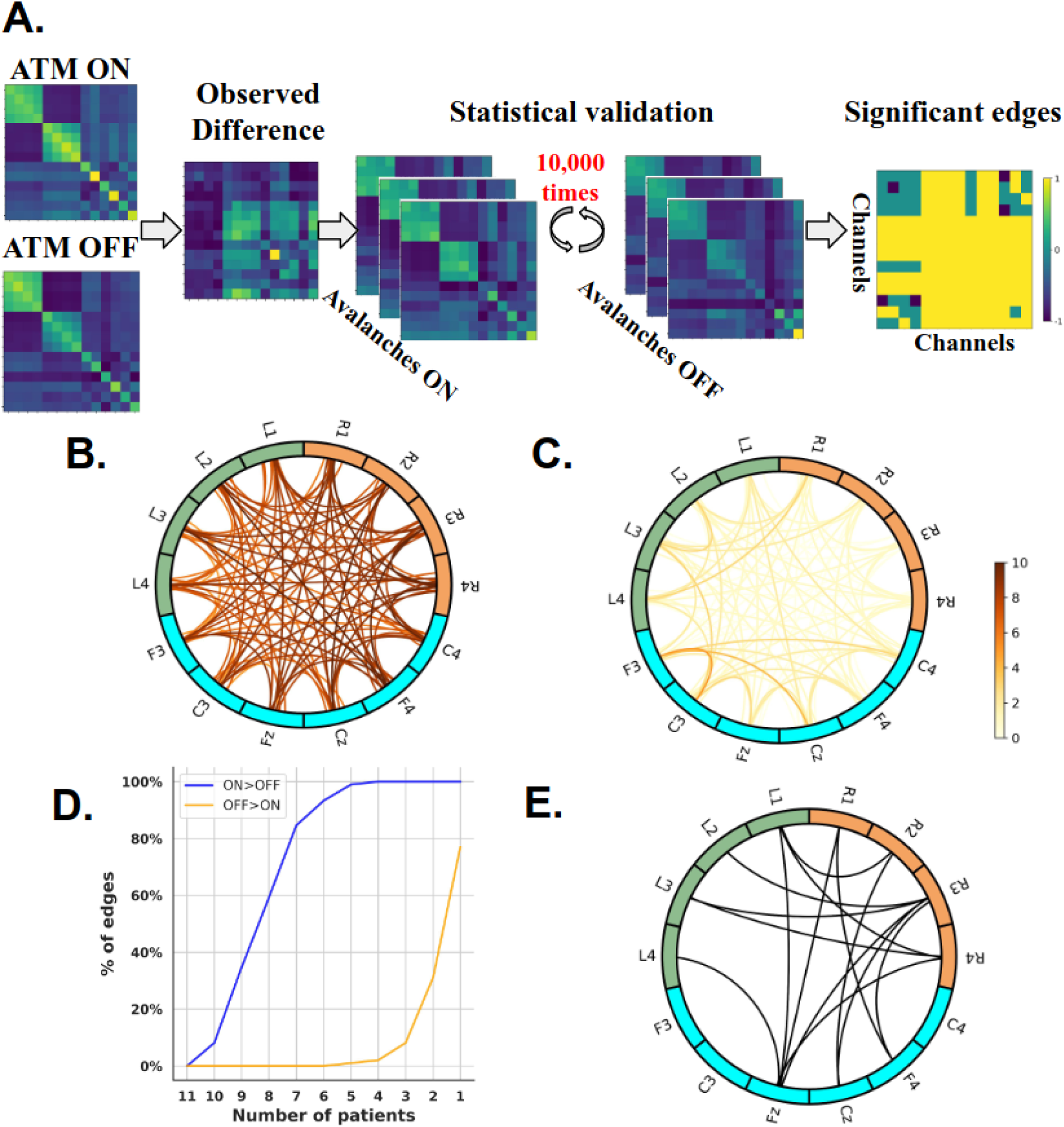
Edge-wise analysis. **(A) Statistical pipeline** used to identify the edges that exhibit significant differences between the ON-levodopa and OFF-levodopa conditions adapted from (Corsi et al., 2023). The ATM_ON_ (ATM in ON-levodopa state) was subtracted, edge-wise, from the ATM_OFF_ (ATM in OFF-levodopa state). The significance is assessed by a permutation test, through iteratively randomizing the labels of avalanche-specific transition matrices (i.e.,ON-levodopa medication and OFF-levodopa medication) 10,000 times, before averaging and computing the differences. The p-values are calculated as the proportion of random differences (separately for ATM_ON_ - ATM_OFF_ and ATM_OFF_ - ATM_ON_) greater than the observed difference for each patient. P-values are considered significant when <0.05 after BH correction for multiple comparisons. **(A) right panel: significance result for one patient**; in this representation, the color code corresponds to the three possible results either ON > OFF (value=1), OFF > ON (value=-1), or non-significant edges (value=0). **(B) Cumulative ON>OFF edge consistency.** The color code reflects the number of significant ON>OFF occurrences across patients. **(C) Cumulative OFF>ON edge consistency.** The color code reflects the number of significant OFF>ON occurrences across patients. **(D) Percentage of edges that are significant for at least n patients, 1≤n≤11 in both cases: the ON-levodopa greater than OFF-levodopa state, and the OFF-levodopa greater than ON-levodopa state. (E) Edge-wise group-level analysis.** The statistical significance of the differences was tested with the Wilcoxon signed-rank test; significant edges (p-values < 0.02) are shown after applying the BH correction for multiple comparisons.

We then compared edge-significance matrices across patients. First, we separated the ON>OFF significant edges from OFF>ON ones, and cumulated them over all patients (see Figures 4.B-C). As expected, a vast majority of edges were statistically significantly stronger in the ON vs. OFF states, but still a subset of patients showed stronger edges in the OFF state, in particular in cortico-cortical edges (see Fig. 4C). Secondly, for each edge, we counted the number of patients for whom it was significantly greater in the ON condition and the number of patients for whom it was greater in the OFF condition. Then, we computed the percentage of edges significantly stronger in the ON/OFF condition than in the OFF/ON condition for at least n patients, with 1**≤**n**≤**11 (see Fig. 4D). From this analysis, we found that all the edges are significantly stronger in the ON condition than in the OFF condition for at least 4 patients. Moreover, we found that more than 40 % of the edges were significantly stronger in the ON than in the OFF condition for at least 9 patients, while specific edges were consistently greater in the OFF vs ON states, but for a maximum of 5 patients.

Interestingly, we found a higher number of consistently significant edges (for more than 9 patients) between the subthalamic nuclei contacts and motor cortex EEG channels (60%) than within the subthalamic nuclei (29 %) or within the motor cortex (11%) in ON-levodopa with respect to OFF-levodopa condition (see Figure 4-1). This suggests that medication affects avalanches globally spreading within and between the STNs and the motor cortices. Furthermore, this higher prevalence of STN-cortical significant edges is consistent with our previous findings, showing that cross-modal edges seem to constitute a reliable indicator of levodopa-induced dynamic changes.

To confirm this analysis, we also computed the edge-wise group-level statistics. Specifically, we evaluated the differences in avalanche transition matrices (ATMs) for each edge across all patients between the ON-levodopa and OFF-levodopa conditions and assessed significance using the Wilcoxon signed-rank test. To correct for multiple comparisons, we applied the BH correction. The group-level analysis was found to be consistent with the subject-level one. In particular, when considering only the edges with a significant ON>OFF difference at the group level (p ≤ 0.02), 60% of them are connecting the STN contacts and the motor cortex EEG channels. Conversely, fewer significant edges are observed within the STN (33%) or within the motor cortex (7%) (see Figure 4.E). This result confirms that the differences in avalanche propagation between medication states more significantly impact the interaction between the STN (and likely other basal ganglia) and the motor cortex.

Incidentally, we observed a clear asymmetry in the distribution of significant edges, with a higher proportion located in the right hemisphere. This asymmetry could be partly explained by the predominance of side-onset disease in the left hemisphere (see Table 1). Thus, the effect of the medication would be more significant contralaterally with respect to the initially affected region, consistent with previous results on the asymmetric effects of levodopa (Martinu et al., 2014).

## Discussion

In the present work, we set out to test the hypothesis that avalanches may be affected, both in terms of statistical features and preferred spatial trajectories, by the administration of levodopa to Parkinsonian patients. This may offer a marker to track the neurophysiological changes induced by medication at the individual level in large-scale brain dynamics. Our findings confirm that neuronal avalanches occur and spread differently depending on the medication condition. In particular, while more avalanches are found in the OFF-levodopa state, they tend to spread more in the ON-levodopa state. Our results also show that the differences in the propagation of avalanches between medication conditions is a global effect, involving interactions within and between STN and the motor cortex, suggesting the altered propagation of neuronal avalanches across these areas as a promising candidate for tracking the neurophysiological changes induced by levodopa at the individual level. Furthermore, we discovered that the increase in the overall propagation of avalanches is associated with motor improvement after levodopa administration. Hence, the L-Dopa-induced changes in the ATMs might be related to the therapeutic response.

Our findings indicate a higher frequency of avalanches in the OFF-levodopa state compared to the ON-levodopa state and, conversely, a higher inter-avalanche interval durations in the ON-state. On the one hand, this finding is reminiscent of the known pathological hypersynchronization that is present in Parkinsonian patients and more evident without therapy (Brown, 2003; Fogelson et al., 2006; Lalo et al., 2008). On the other hand, we found a reduced number of avalanches, lasting longer and being larger in size in the ON-levodopa state. This suggests that the picture might be more complex than previously thought, with changes in the dynamical structure hardly reported using a conventional oscillatory perspective.

Despite the lower number of neuronal avalanches in the ON-levodopa state, their spreading, once they occur, appears to be facilitated by the presence of levodopa, as evident from the generally higher transition probabilities in the ON state. This result demonstrates that neuronal avalanches are more likely to spontaneously propagate in the ON-levodopa condition for Parkinsonian patients. Conversely, while more avalanches start during the OFF state, they fail to effectively recruit a high number of brain areas. Furthermore, we show that on closer topographical analysis, the differences between the conditions are not equally present throughout the brain but, rather, they mainly involve the interaction between the STN and the motor cortex in the sense of more spreading of avalanches taking place under levodopa.

Our results align with previous neuroimaging research indicating that in Parkinson’s disease, dopaminergic depletion is linked with malfunctioning of the basal ganglia motor circuit (BGMC) (Haslinger et al. 2001; Wu et Hallett 2005; Herz et al. 2014; Gao et al. 2017), along with reduced connectivity of the striato-thalamo-cortical motor pathways (Wu et al., 2011b; Sharman et al., 2013; Michely et al., 2015). In addition, prior neuroimaging research has indicated that the administration of levodopa might partially restore the abnormal functional connectivity (FC) in the BGMC circuit among patients with Parkinson’s disease. For instance, it may achieve this by boosting neural activity within the supplementary motor area and striatum (Haslinger et al., 2001; Buhmann et al., 2003; Kraft et al., 2009; Palmer et al., 2009; Wu et al., 2009a; Martinu et al., 2012) and reestablishing connectivity in the striato-cortical motor pathway (Wu et al., 2009b; Agosta et al., 2014a). The restoration of striatal-cortical connectivity may result in restoration of cortico-subthalamic connectivity as observed in our results via the indirect pathway.

Furthermore, we demonstrated that the increase in the overall propagation of avalanches correlates with motor improvement after levodopa administration. This suggests that the proposed pipeline, beyond tracking neurophysiological changes related to the administration of levodopa, might provide a readout to the clinical improvement of the patients. This is aligned with previous studies, which showed a correlation between changes in FC and improvements in clinical symptoms following levodopa treatment (Wu et al., 2009c, 2011a; Agosta et al., 2014b; Gao et al., 2017; Shen et al., 2020). In addition, the correlation between the ratio of the average ATMs in the ON and the OFF states with the clinical improvement after levodopa administration reached significance when computed selectively on the STN-cortex edges. This might indicate that clinical improvement relates the most to the coordination of activities between the STN and the motor cortex. These results complement the evidence provided above about the differences in avalanche propagation between medication states.

Of note, the reported differences of the propagation of avalanches are observed not only at the subcortical level through STN activity but also at the cortical level through EEG activity, which could be relevant, for example, for clinical purposes, given the ease of access to EEG data. We also found a significant increase in the propagation of avalanches for both short-range (within the subthalamic nuclei and within the motor cortex) and long-range (between the subthalamic nuclei and the motor cortex) connections in the ON-levodopa state compared to OFF-levodopa states. Also, this means that the effects can be observed within a modality as well as between modalities. To our knowledge, there are no previous works that have studied the probability of propagation of avalanches across modalities.

In summary, our study revealed that the administration of Levodopa enhanced the ease of propagation of neuronal avalanches between the motor cortex and the STN. This aligns with prior studies suggesting that treatment using levodopa partially restores the function of the basal ganglia motor circuit (Haslinger et al., 2001; Buhmann et al., 2003; Kraft et al., 2009; Palmer et al., 2009; Wu et al., 2009a; Martinu et al., 2012; Gao et al., 2017; Shen et al., 2020). Additionally, we found that the increase in the overall propagation of avalanches is correlated with motor improvement after levodopa administration. Therefore, proposing the ATM as a potential biomarker serves not only to track the medication’s effect but also to track the individual clinical improvement in Parkinson’s disease. Furthermore, and more generally, our study adds to the growing literature showing that conceptualizing brain dynamics in PD in terms of aperiodic bursts yields relevant information that might not be apparent in a more oscillatory perspective. Therefore, our findings provide insights into the neural mechanisms underlying the effect of levodopa therapy at the whole-brain level in Parkinson’s disease.

## Data availability & code accessibility

The codes employed in this study are available at https://github.com/Hasnae12/neuronal_avalanches_PD. However, due to the clinical nature of the data, it can’t be publicly available.

## Extended Data

**Table1.** Clinical features of some patients. F = female; M = male; L = left; R = right; AR=Akinetic rigid; UPDRS= Unified Parkinson’s Disease Rating Scale. Clinical data from 10 PD patients shows a reduction in the UPDRS after the intake of medication, indicating a positive response to medication.

| Patient | Age Range (years) | Sex | Disease Duration (years) | First symptom | Side onset | UPDRS III OFF | UPDRS III ON | Delay between surgery and recordings (days) | Medication |
| --- | --- | --- | --- | --- | --- | --- | --- | --- | --- |
| P01 | 51-55 | M | 14 | Tremor | R | 27 | 5 | 3 | ropinirole 8mg; l-dopa 1250mg/j ; entacapone ; rasagiline |
| P02 | 66-70 | F | 12 | AR | R | 31 | 10 | 3 | ropinirole 8mg; l-dopa 700mg/j ; entacapone |
| P03 | 66-70 | M | 10 | Tremor | R | 32 | 5 | 3 | piribedil 150mg/j ; l-dopa 1250mg/j ; entacapone |
| P04 | 51-55 | F | 8 | Tremor | L | 40 | 16 | 3 |  |
| P05 | 51-55 | F | 10 | AR | L | 11 | 2 | 3 | ropinirole 16mg ; l-dopa 400mg |
| P06 | 61-65 | F | 12 | AR | L | 21 | 5 | 3 | ropinirole 20mg; ldopa 725mg; entacapone; rasagiline 1mg |
| P07 | 56-60 | M | 17 | AR | R | 27 | 2 | 3 | pramipexole 2,10mg; ldopa 1200mg; entacapone |
| P09 | 61-65 | M | 14 | Tremor | L | 29 | 9 | 3 | piribedil 150mg; ldopa 900mg; entacapone |
| P10 | 61-65 | M | 7 | Tremor | L | 31 | 5 | 3 | ropinirole 24mg; ldopa 800mg; entacapone; rasagiline 1mg |
| P11 | 56-60 | F | 8 | Tremor | L | 20 | 8 | 5 | piribedil 150mg; ldopa 1000mg; entacapone |

**Figure 1-1.**
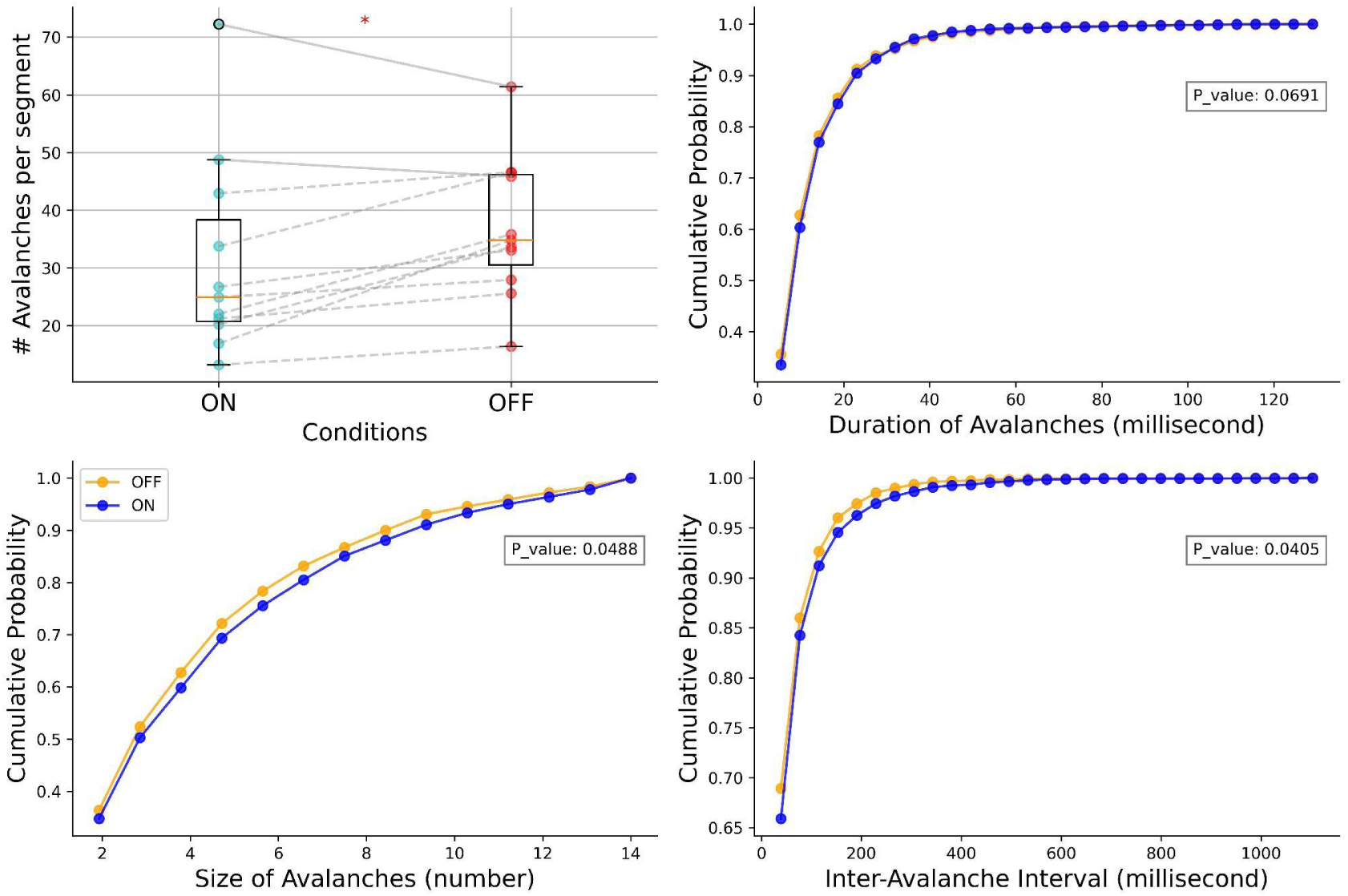
Group level analysis of avalanches features. In **(A)** We compared the number of avalanches per segment in ON-levodopa versus OFF-levodopa states at the group level. The significance of the difference was assessed using the paired t-test, resulting in a p-value of 0.041. We also compared the cumulative distribution function at the group level for duration **(B)**, size **(C)**, and inter-avalanche interval **(D)**. We tested the significance of the results using the Kolmogorov-Smirnov (K-S) test, resulting in p_values of 0.069 for duration, 0.048 for size, and 0.040 for Inter-avalanche interval.

**Figure 1-2.**
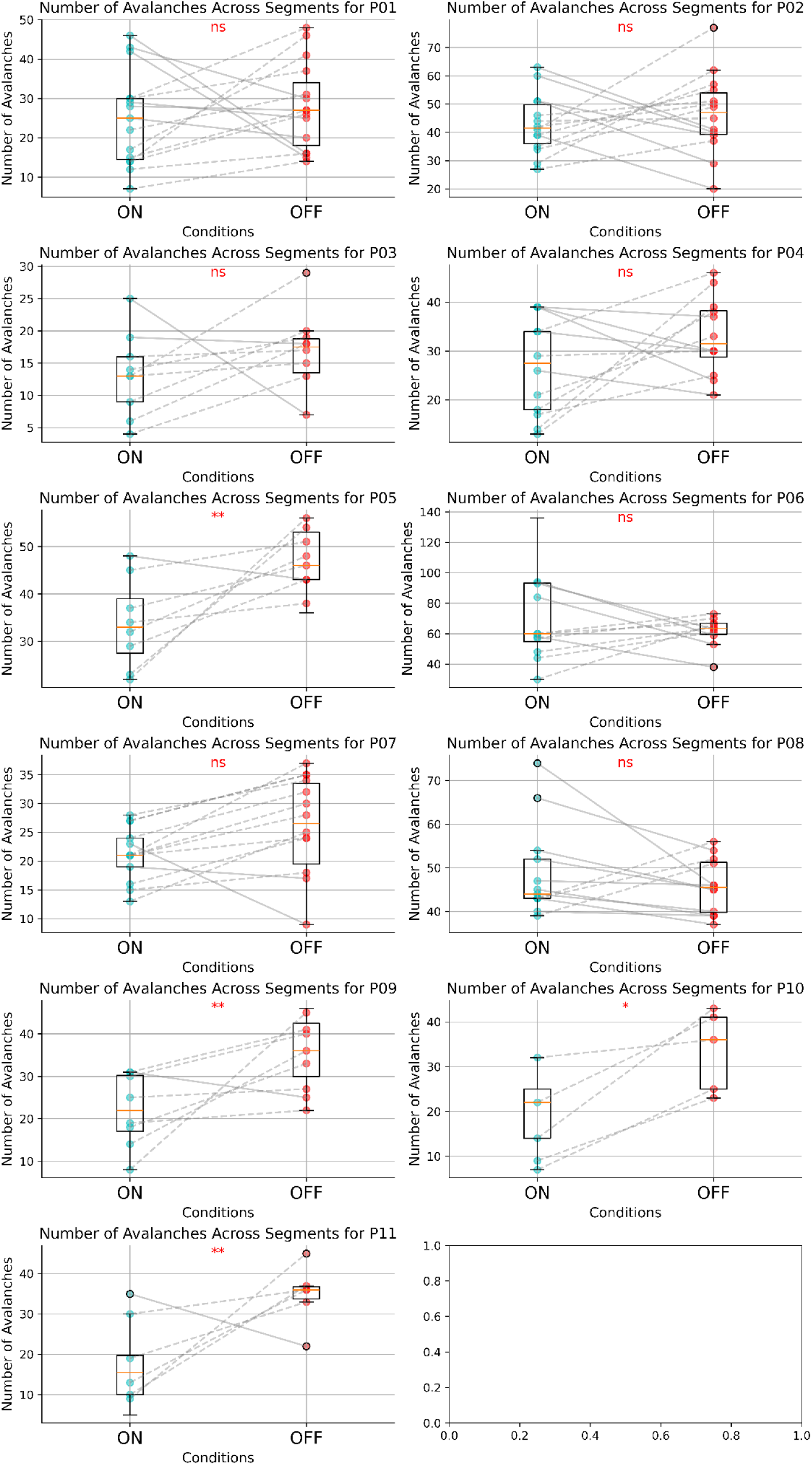
Subject level analysis of avalanches features. Comparison of the number of avalanches across segments for each patient in the ON-state and OFF-state, along with Mann-Whitney U test results. Due to the different number of segments between ON and OFF conditions per patient, we plotted the number of avalanches across segments, using the minimum number of segments between conditions for each patient. The significance level are indicated as *** : p< 0.001, ** : p<0.01, *: p<0.05, ns = non-significant.

**Figure 1-3.**
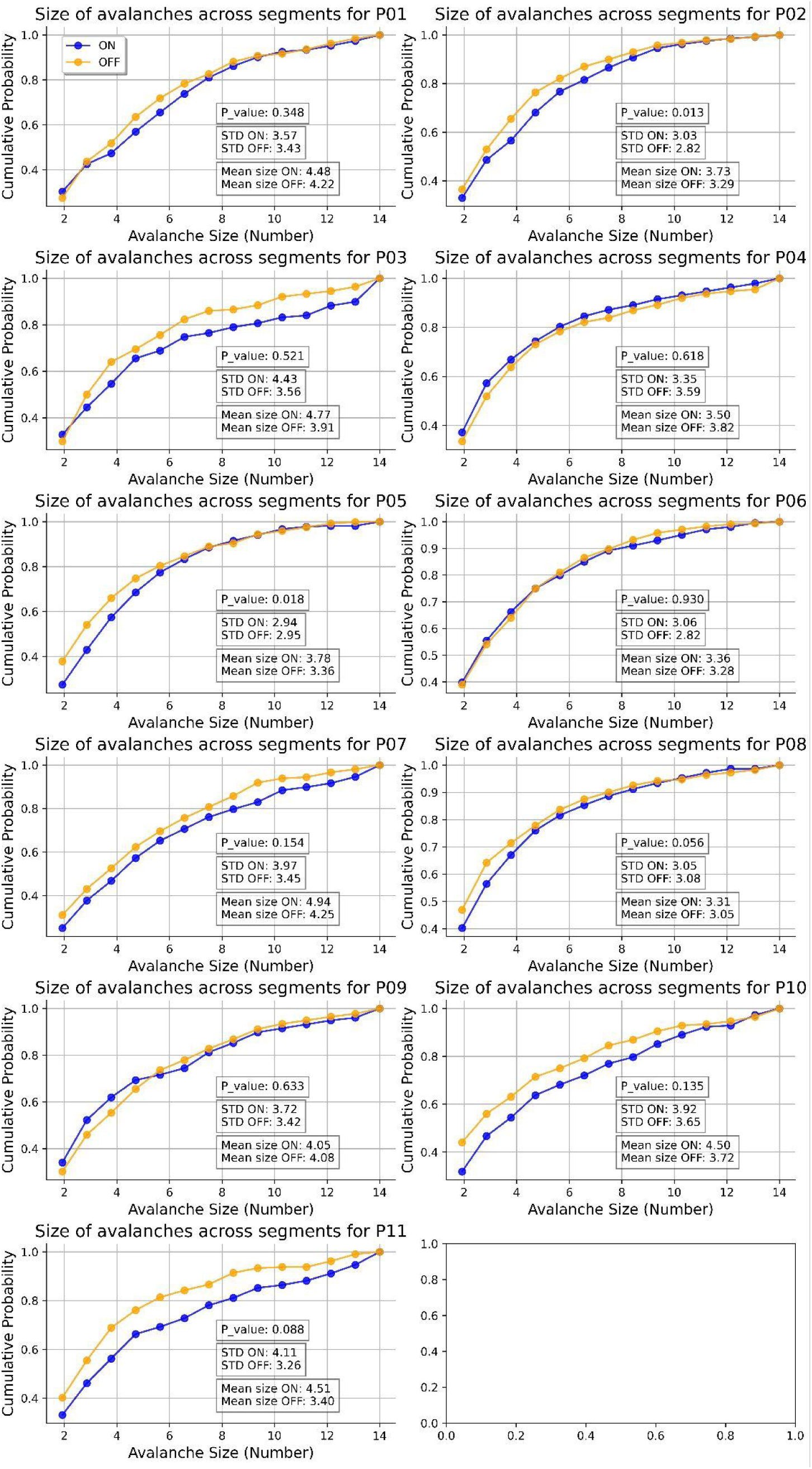
Subject level analysis of avalanches features. Comparison of the cumulative distribution functions of avalanche sizes across segments for each patient in the ON-state and OFF-state, along with Kolmogorov-Smirnov (K-S) test results, mean values, and standard deviations.

**Figure 1-4.**
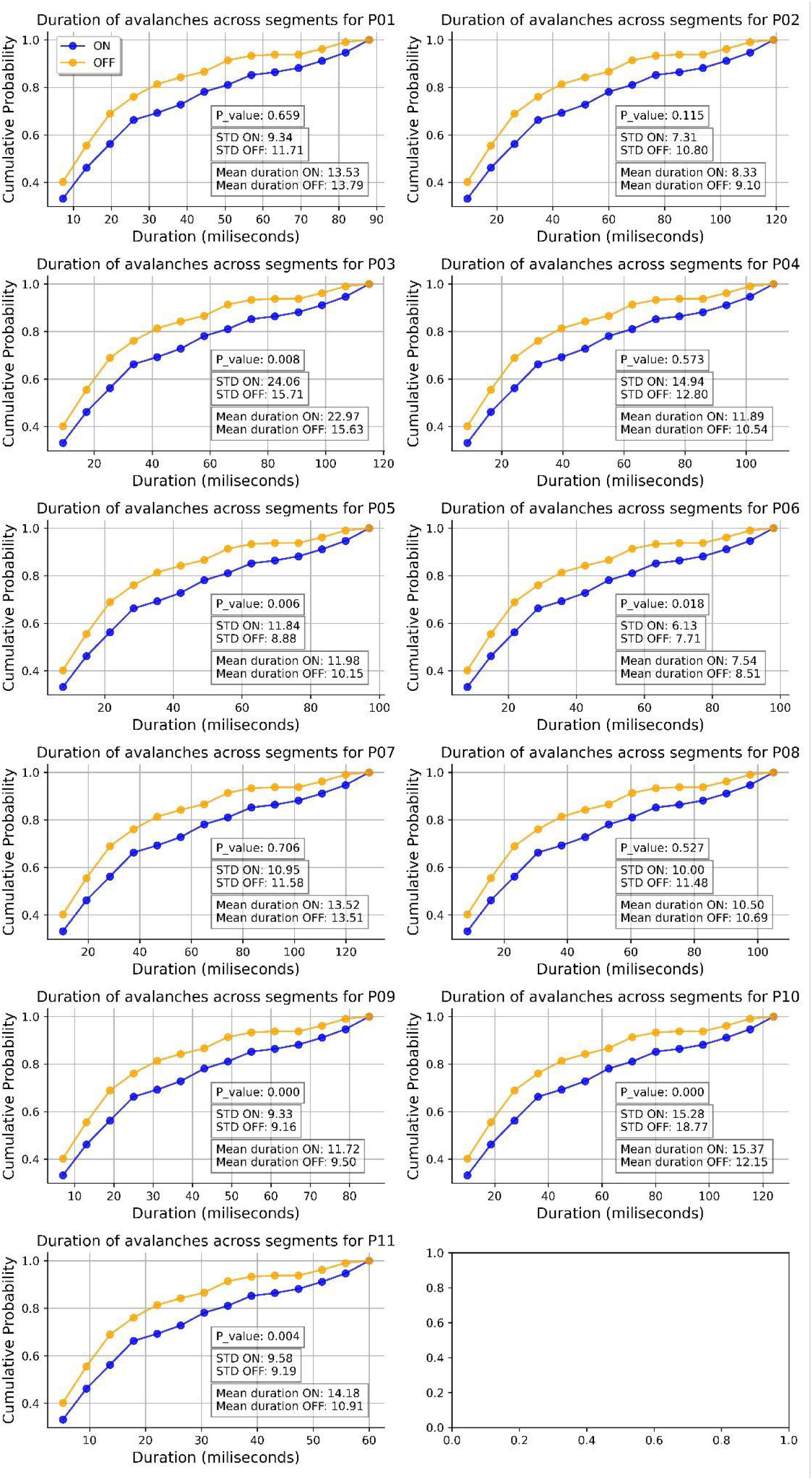
Subject level analysis of avalanches features. Comparison of the cumulative distribution functions of avalanche durations across segments for each patient in the ON-state and OFF-state, along with Kolmogorov-Smirnov (K-S) test results, mean values, and standard deviations.

**Figure 1-5.**
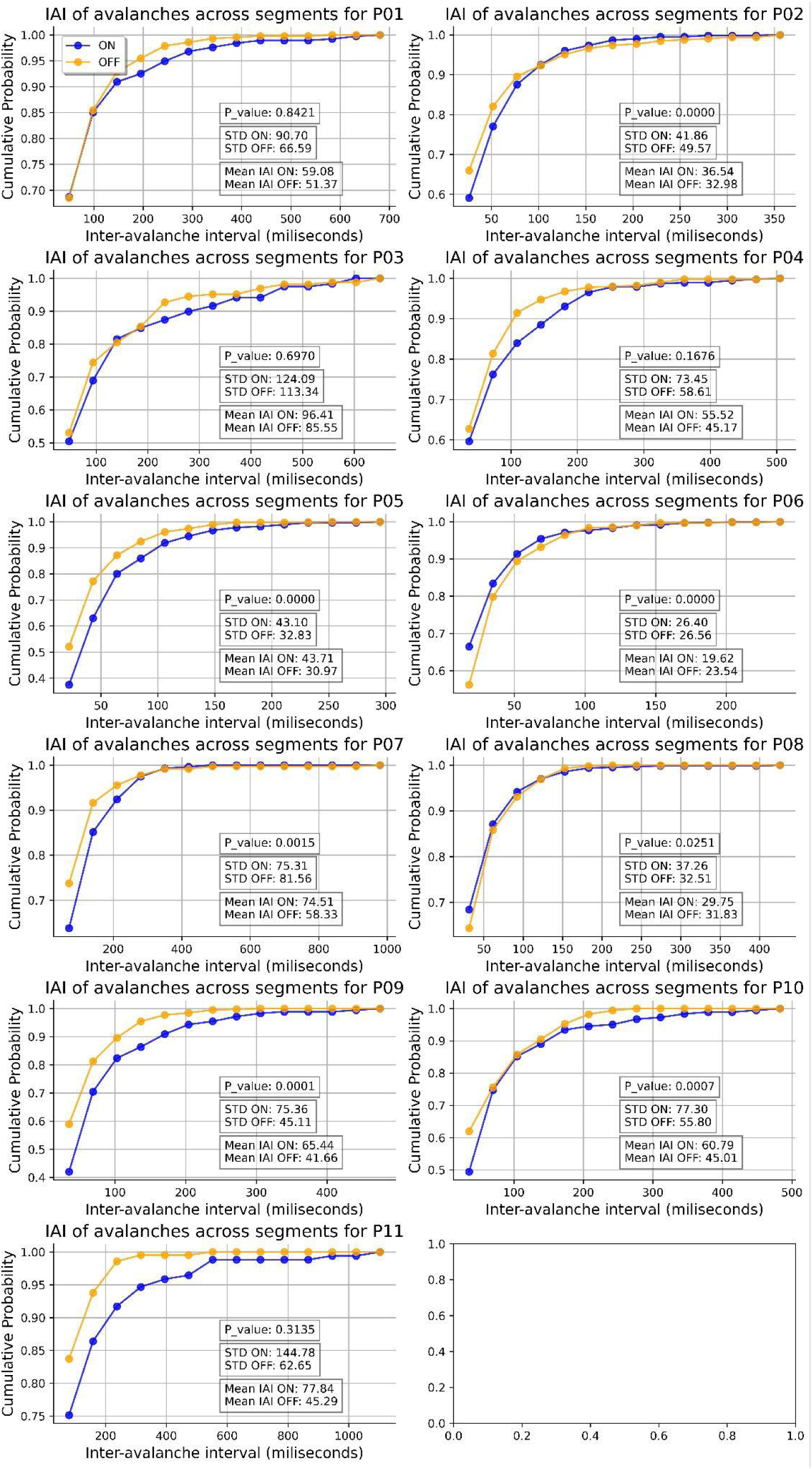
Subject level analysis of avalanches features. Comparison of the cumulative distribution functions of avalanche Inter-avalanche intervals (IAI) across segments for each patient in the ON-state and OFF-state, along with Kolmogorov-Smirnov (K-S) test results, mean values, and standard deviations.

**Figure 2-1.**
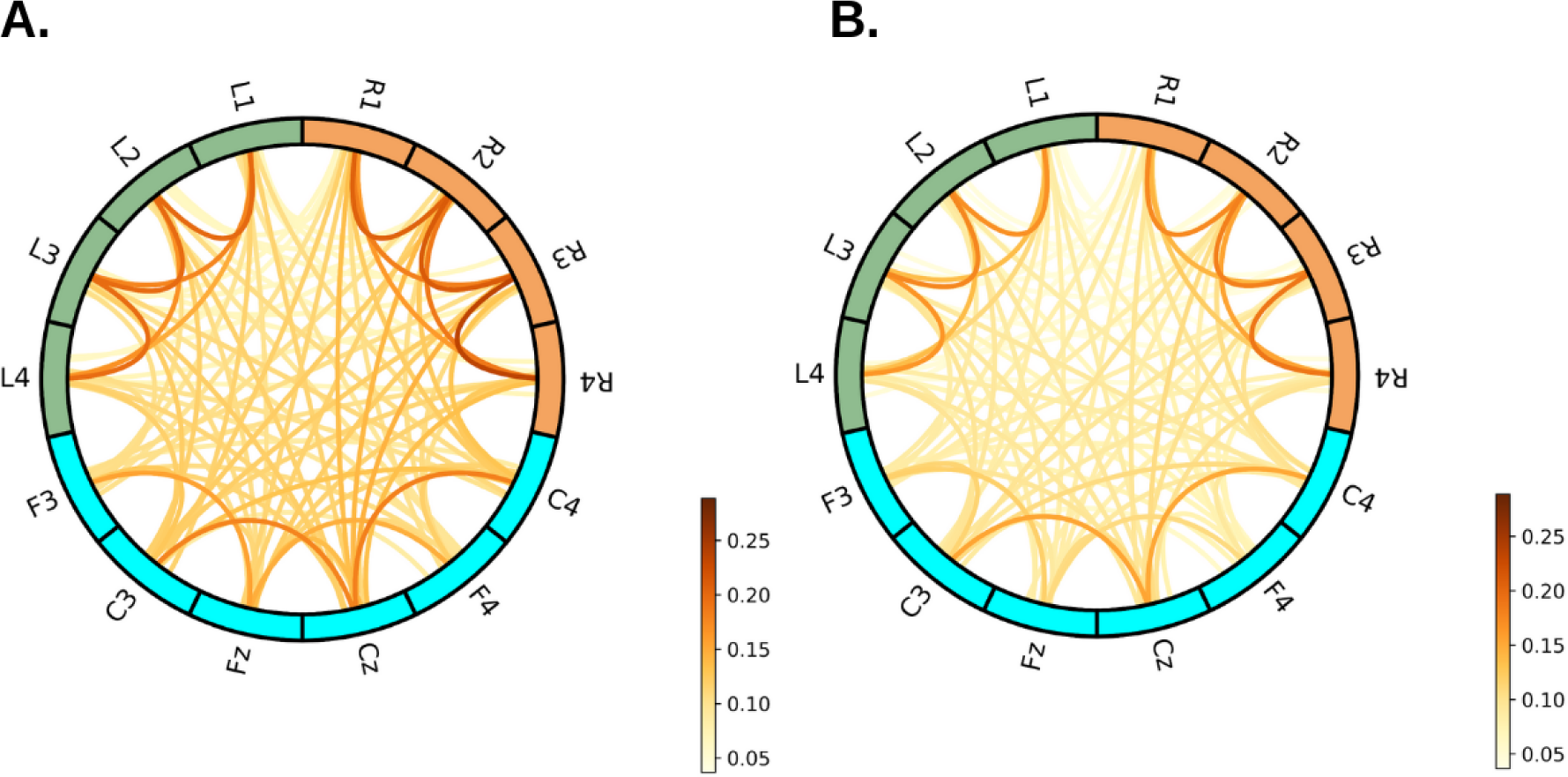
Average of the original Avalanche Transition Matrices across patients in ON-levodopa and OFF-levodopa conditions. In **(A),** we plotted the average ATMs in the ON-levodopa across all patients. In **(B),** we plotted the average ATMs in the OFF-levodopa across all patients. We represented the brain as a network, where channels are nodes linked by edges. For the nodes, the green represents the left LFP contacts, orange represents the right LFP contacts, and blue represents the EEG channels.

**Figure 4-1.**
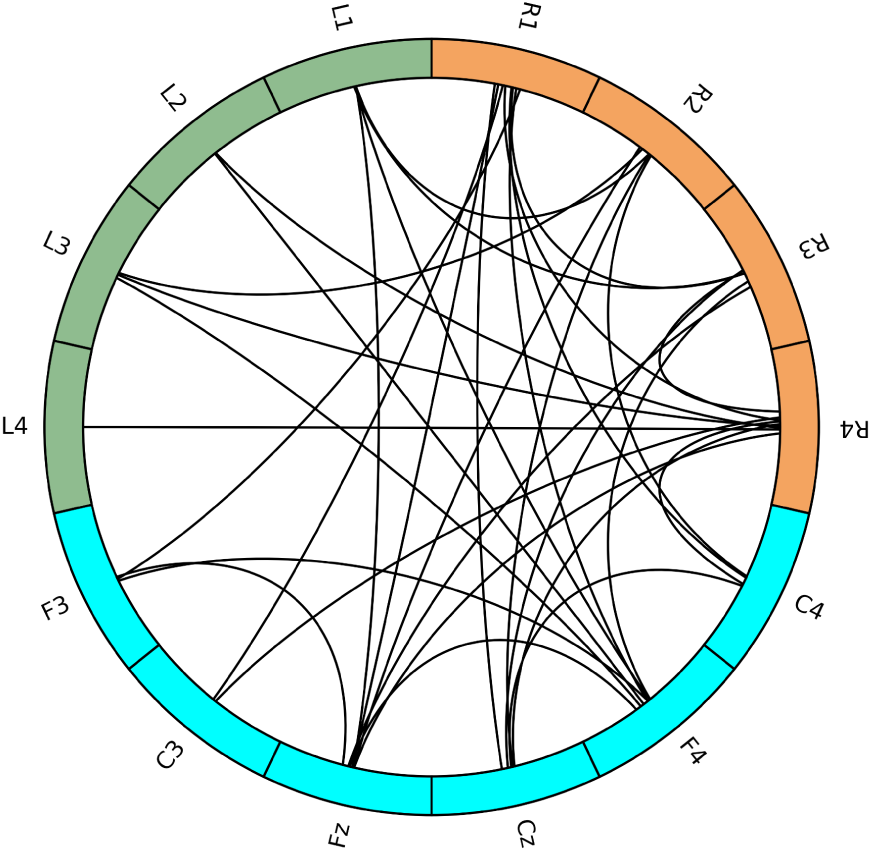
Edges with cumulative consistency >= 9. patients in the ON>OFF case

